# How urgent do intravitreal anti-VEGF injections need to be to justify the risk of transmitting COVID-19? Proof-of-concept calculations to determine the Health Adjusted Life-Year (HALY) trade-off

**DOI:** 10.1101/2020.04.27.20075085

**Authors:** Matt James Boyd, Daniel Andrew Richard Scott, David Michael Squirrell, Graham Ashley Wilson

## Abstract

**Background:** Clinical ophthalmological guidelines encourage the assessment of potential benefits and harms when deciding whether to perform elective ophthalmology procedures during the COVID-19 pandemic, in order to minimize the risk of disease transmission.

**Method:** We performed probability calculations to estimate COVID-19 infection status and likelihood of disease transmission among neovascular age-related macular degeneration patients and health care workers during anti-VEGF procedures, at various community prevalence levels of COVID-19. We then applied the expected burden of COVID-19 illness and death expressed through health-adjusted life-years (HALYs) lost. We compared these results to the expected disease burden of severe visual impairment if sight protecting anti-VEGF injections were not performed.

**Results:** Our calculations suggest a wide range of contexts where the benefits of treatment to prevent progression to severe visual impairment or blindness are greater than the expected harms to the patient and immediate health care team due to COVID-19. For example, with appropriate protective equipment the benefits of treatment outweigh harms when the chance of progression to severe visual impairment is >0.044% for all scenarios where COVID-19 prevalence was one per thousand, even when the attack rate in the clinical setting is very high (5-43%).

**Conclusion:** Unless COVID-19 prevalence is very high, the reduced disease burden from avoiding visual impairment outweighs the expected HALYs lost from COVID-19 transmission. This finding is driven by the fact that HALYs lost when someone suffers severe visual impairment for 5 years are equivalent to nearly 400 moderate cases of infectious disease lasting 2 weeks each.

## Introduction

The coronavirus causing COVID-19 disease began circulating in Wuhan, China, in November 2019, quickly becoming a global pandemic. Italy was an early epicenter of the pandemic and Italy’s national medical federation reported that 10% of responding health-care workers had been infected with 125 attributed-physician deaths recorded from 11 March to 17 April 2020.^1^ In New York City when that city was the epicenter of the pandemic (March–April 2020), ophthalmology residents had the fifth highest proportion of infections among specialties, with 5% of residents confirmed to have acquired COVID-19. It cannot be ascertained whether this was in the line of work or not. For comparison 2.4% of GPs and 1.1% of diagnostic radiologists had confirmed infection.^2^ The risk of transmission from patient to health care team, and among members of health care teams, clearly must be mitigated.

Advice in recent ophthalmologic guidelines suggests that a clinical evaluation of the benefits and harms of performing eye procedures during the COVID-19 pandemic should determine whether to proceed or defer interventions.^3^ This means that clinicians should be using clinical judgment to weigh the probability of serious progression of eye disease against the probability that COVID-19 might spread among patients and health care staff. Patients with non-urgent clinical conditions (such as stable glaucoma and amblyopia) and non-urgent surgical conditions (such as cataract or epiretinal membrane) can be scheduled later.^3^ For those requiring urgent clinical assessment or procedures, stringent isolation and protection measures should be integrated into the workflow. One major consideration is the availability of personal protective equipment (PPE) for health care workers.^4^ However, it is useful to have an approximate quantitative estimate of benefits and harms of proceeding with an intervention to guide clinical decision-making.

In the present analysis we focus on intravitreal injections. These have become the most common procedure in both ophthalmology and medicine.^5^ Injection of anti-vascular endothelial growth factor (anti-VEGF) for treatment of neovascular age-related macular degeneration (nAMD) is the most common indication.^6^ Since their introduction to our setting of New Zealand there has been a significant reduction to nAMD-associated blindness.^7^ The expected cost per quality adjusted life-year for sight improvement/maintenance from this treatment is estimated at NZ$2,900. In New Zealand, most nAMD treatments are delivered by the public health service, with less than 10% of volume performed by private healthcare providers.^7^

Triage guidelines released by The Royal Australian and New Zealand College of Ophthalmologists (RANZCO) recommend continuing clinic appointments during the COVID-19 pandemic for nAMD patients with a high urgency.^8^ The American Macular Degeneration Foundation shares a similar position, stating that ‘anti-VEGF injections are essential for those who require them’.^9^ This is because the risks of stopping or delaying treatments for nAMD are significant. A delay in providing treatment is known to increase the risk of disease reactivation and visual loss.^10^ As a lifelong chronic disease, even with good disease control, continued anti-VEGF injections are recommended to reduce the risk of reactivation.^11^ We know from an observational study of 434 eyes where treatment was suspended, that 41% experienced disease reactivation within the first year.^12^ In a large cohort study from the United Kingdom (UK), just 6.9% of nAMD patients became legally blind following 2 years of anti-VEGF, compared to a predicted figure of 15.7% for untreated patients.^13^ Furthermore, a modelling simulation study in the UK using data from a COVID-19 lockdown period, estimated a 50% increase in the number of patients with severe visual impairment from a 3-month treatment delay.^14^

COVID-19 imposes a high burden of morbidity and mortality upon communities through illness and death. However, severe vision impairment and blindness due to untreated eye conditions such as AMD also imposes quality of life losses. The clinical judgment equation must determine whether to treat or defer after balancing these competing impacts on health.

In this paper, we compare the burden of disease due to severe vision impairment against the possible burden due to COVID-19 transmission among nAMD patients and the healthcare team at various community levels of infection prevalence. We aimed to estimate the probability of progression to severe vision impairment in the better seeing eye, that might justify nAMD treatment rather than deferral for individual cases during the COVID-19 pandemic.

## Method

We developed a simple model of COVID-19 transmission and illness impacts. This model accounts for the true background prevalence of COVID-19 disease in the community, and therefore the probability that health care workers or the patient (drawn randomly from the population) are currently infected, the probability of transmission among these individuals at the time of treatment based on attack rate data, the health impact in terms of vision impairment, COVID-19 infection, and life years lost due to death from COVID-19. The parameters used in the model are displayed in Table 1.

**Table 1:**
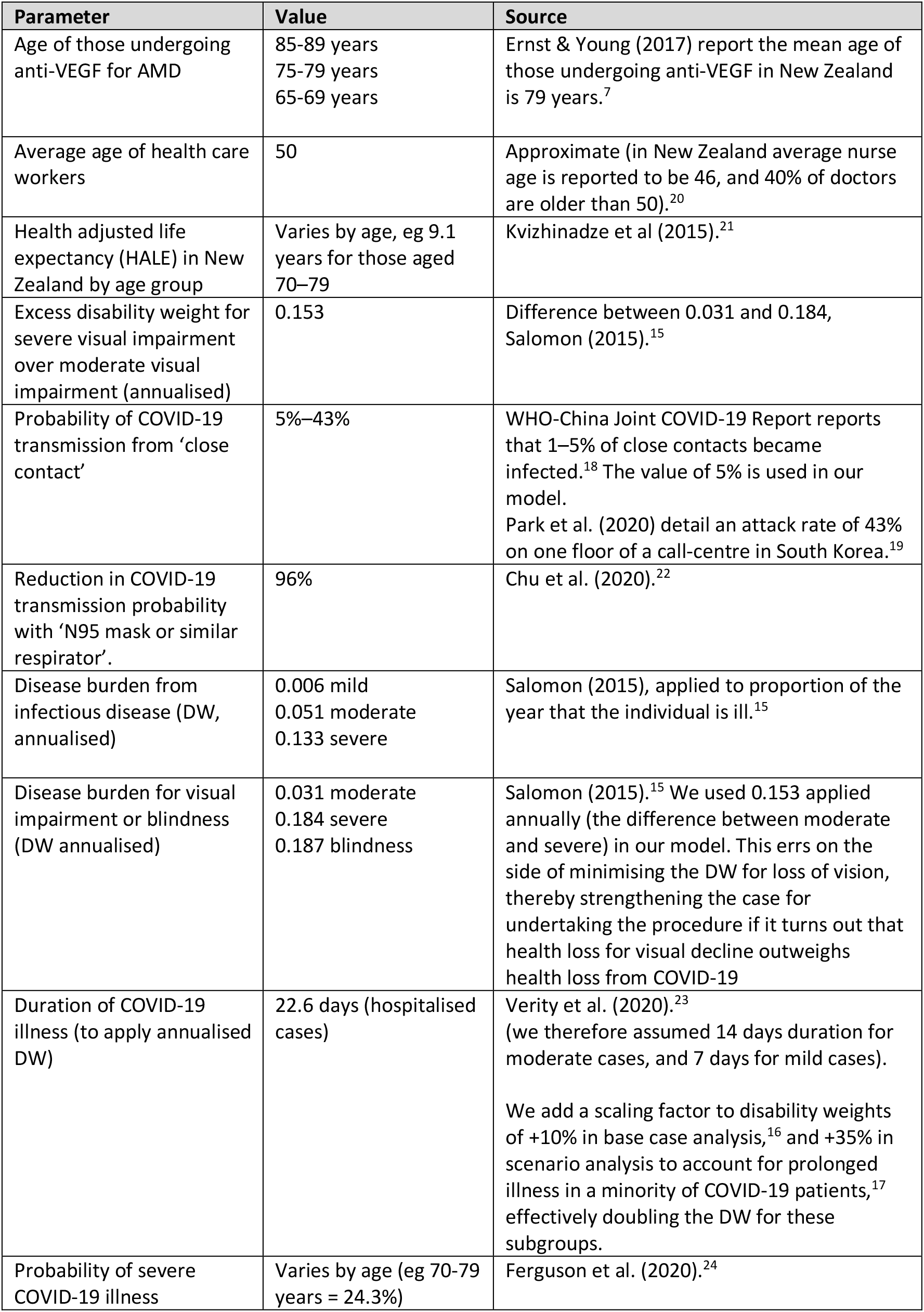
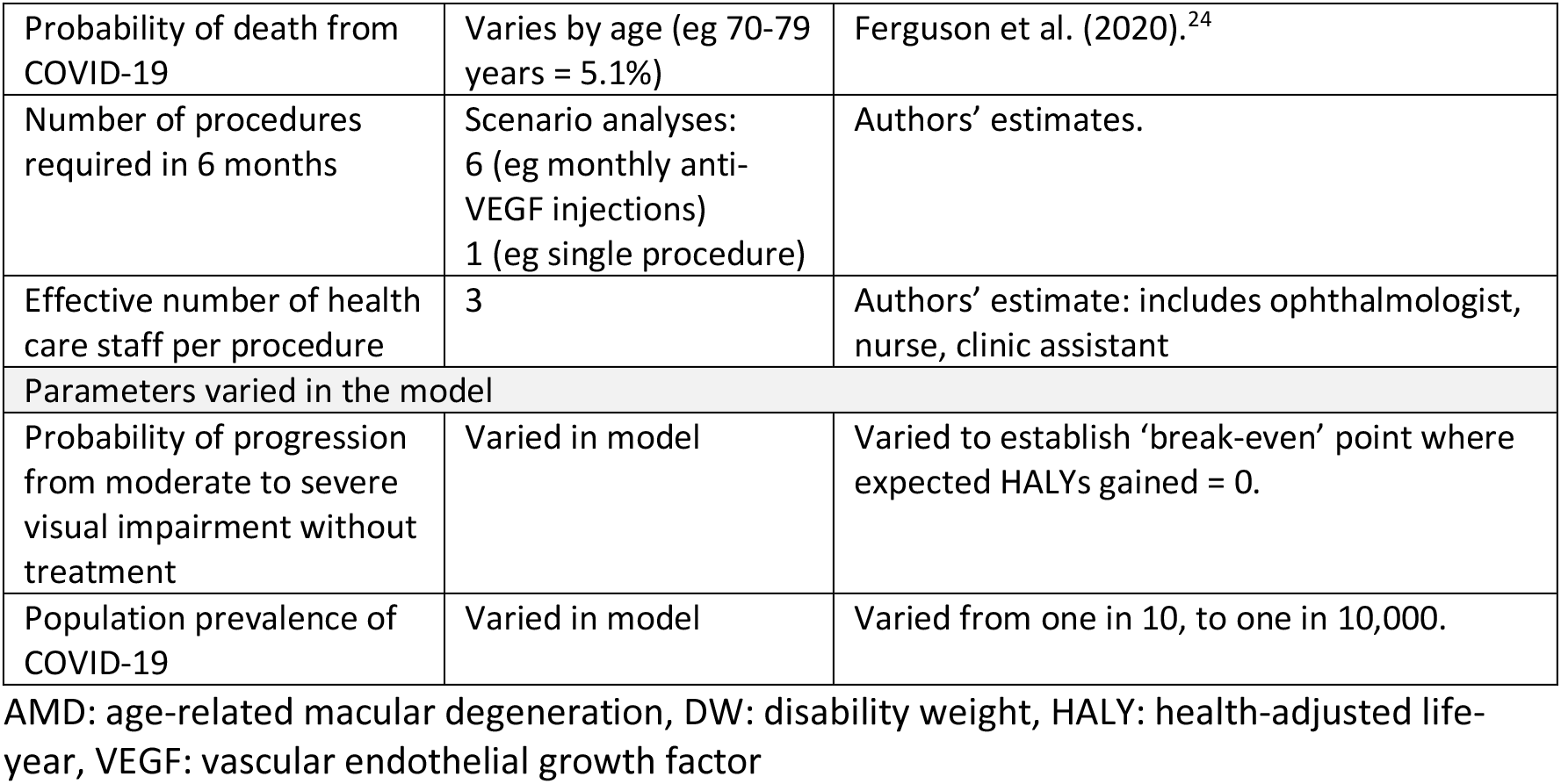
Parameters used in the proof-of-concept model to estimate expected health-adjusted life-year impact of proceeding with anti-VEGF injections in the context of COVID-19 pandemic.

The health impact of COVID-19 and visual impairment was compared using calculations of health-adjusted life years (HALY), which measure the combined effects of mortality and morbidity. One HALY is equivalent to one year of life lived in good health. For individual diseases, annualized disability weights (DW) have been calculated which when applied for the duration of illness have the net effect of reducing the health of a life-year by a fractional component. The Global Burden of Disease (GBD) Study used surveys to obtain data on paired comparisons of different health states and calculated and published DWs for a range of conditions.^15^ For example, the GBD DW for a ‘moderate infectious disease’ was 0.051, which can be applied for the duration of the disease to quantify a one-time health impact. We note that some people with COVID-19 illness experience lingering symptoms. Data from a UK symptom app suggested this might be 10% of patients.^16^ Data from a US study of those testing positive indicated 35% of people have symptoms beyond 1-2 weeks.^17^ We account for this in the model parameters (see Table 1).

For a chronic condition like severe distance vision impairment (bilaterally) the GBD DW of 0.184 would be applied per annum (for remaining expected lifetime if permanent). In our model life years were health adjusted, meaning that any loss of life years was already adjusted for morbidity such as the expected level of pre-existing illness by age group.

Data from contact tracing in China suggest a risk of infection by COVID-19 to close contacts of 1-5%.^18^ Whereas data from a study of transmission in a South Korean call center suggest an attack rate of up to 43% for those who spend day after day in close proximity without protective equipment.^19^ We modelled both these scenarios, which likely bound any healthcare setting attack rate.

For simplicity we assumed a healthcare team of 3 members for a standard ophthalmologic procedure such as anti-VEGF injection for nAMD (injector, nurse, and an additional assistant/clinic staff). This means the patient experiences three possible exposures to COVID-19 and each healthcare team member is exposed to two other staff and the patient. When calculating the impact of vision loss we assumed the patient already had moderate distance vision loss (better eye), with visual impairment: <6/18 to 6/60, (DW 0.031) and was at risk of progressing to severe distance vision loss, <6/60 to 3/60, or blindness, <3/60, (DW 0.184 and 0.187 respectively) which were treated as equivalently bad outcomes for simplicity. Hence our analysis is one of better seeing eye situations.

Given that severity of illness and death from COVID-19 vary by age, we assumed an average healthcare worker age in New Zealand of approximately 50 years.^20^ We analysed a number of scenarios including where a patients of age 65, 75 and 85 receive monthly anti-VEGFs across six months, also a scenario where a patient of 75 years receives a single treatment. We also evaluated scenarios assuming the transmission probability reported for ‘close contacts’ was reduced by 96% due to the impact of full PPE used by all staff (gloves, goggles, face shields, water resistant gowns, and respiratory protective equipment).

Our coarse-grained model assumed a new health care team at each treatment occasion as a simplifying assumption in order to avoid the complication of tracking those who had been infected and recovered. Population prevalence of COVID-19 was assumed to be static across time for repeat procedures.

Figure 1 illustrates how the model works by calculating the probability that transmission of COVID-19 occurs during treatment and then attributing health impacts to these cases according to age and statistical severity of illness. It also attributes health impacts for progression of visual impairment, and these values are then used to determine the probability of progression which equals the health impact of COVID-19 transmission.

**Figure 1:**
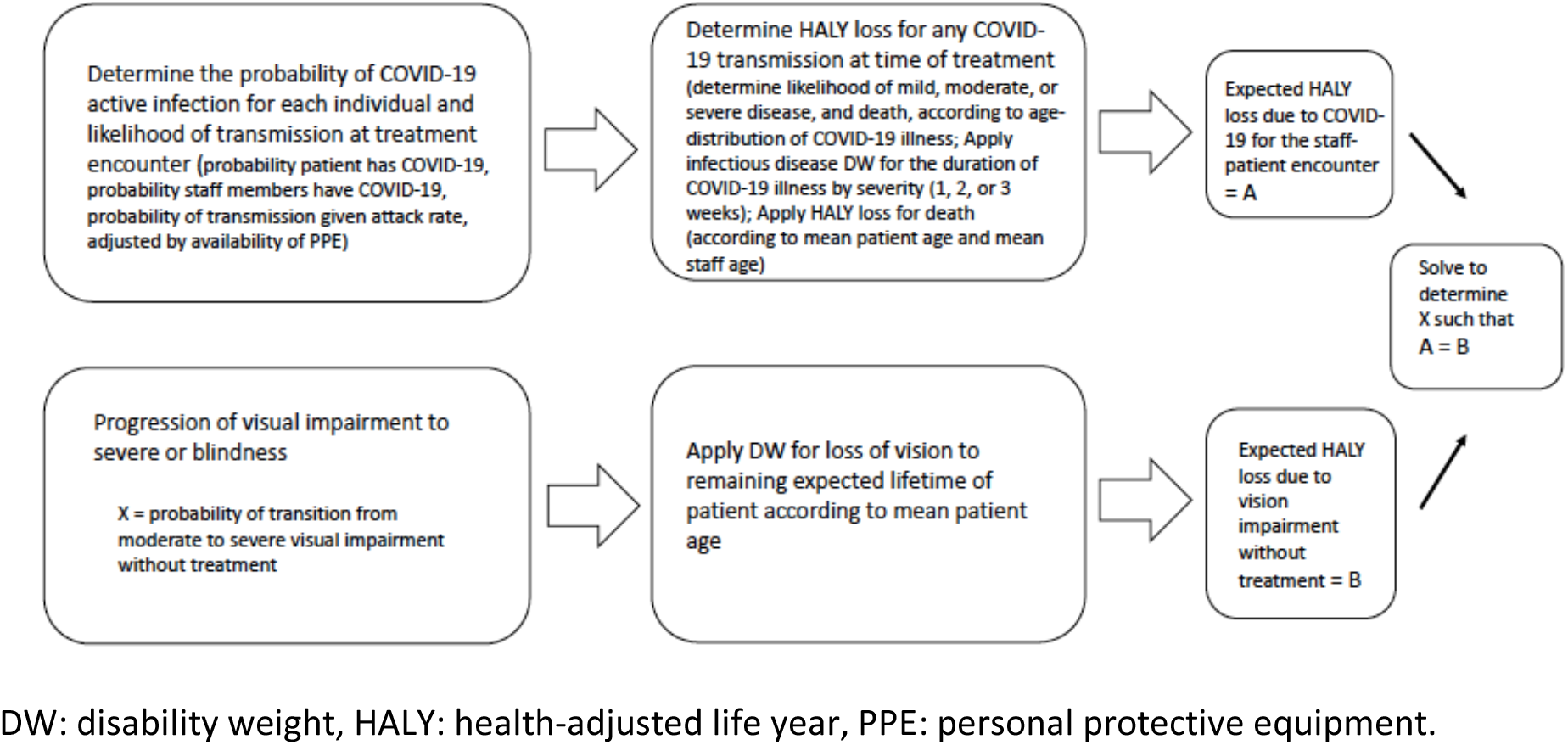
Model to compare likely HALY loss due to COVID-19 transmission among patient and immediate healthcare team against HALY loss due to progression of visual impairment without treatment

## Results

Key results are summarized in Table 2. The results are presented with respect to three dimensions, (1) the ‘optimistic’ presumed values where relevant attack rate of COVID-19 in a clinical setting and additional DW due to lingering symptoms are 5% and +10% respectively, (2) the ‘pessimistic’ presumed values for these two variables (43% and +35%) alongside these and (3) by the community prevalence of active COVID-19 indicated by row headings. The results are expressed as the threshold risk of untreated disease progression necessary for the risk of withholding treatment to be greater than the risk of giving it.

**Table 2:** Threshold risk of untreated disease progression necessary for the risk of withholding treatment to be greater than the risk of giving it: personal protective equipment is available and provides 96% reduction in transmission

| Active community COVID-19 case prevalence | 65-year-old, monthly anti-VEGF injections |  | 75-year-old, monthly anti-VEGF injections |  | 75-year-old, monthly anti-VEGF injections |  | One-time anti-VEGF injection: 75-year-old |
| --- | --- | --- | --- | --- | --- | --- | --- |
|  | optimistic assumptions | pessimistic assumptions | optimistic assumptions | pessimistic assumptions | optimistic assumptions | pessimistic assumptions | pessimistic assumptions |
| 1/10,000 | <0.001% | 0.001% | <0.001% | 0.002% | <0.001% | 0.004% | <0.001% |
| 1/1,000 | 0.001% | 0.011% | 0.003% | 0.022% | 0.005% | 0.044% | 0.004% |
| 1/100 | 0.013% | 0.111% | 0.025% | 0.217% | 0.051% | 0.436% | 0.036% |
| 1/10 | 0.127% | 1.11% | 0.252% | 2.17% | 0.506% | 4.36% | 0.362% |

Table 3 provides the same analysis as Table 2, but in the context where appropriate personal protective equipment is not available and hence the 96% reduction on the attack rate is removed.

**Table 3:** Threshold risk of untreated disease progression necessary for the risk of withholding treatment to be greater than the risk of giving it: appropriate personal protective equipment is not available

| Active community COVID-19 case prevalence | 65-year-old, monthly anti-VEGF injections |  | 75-year-old, monthly anti-VEGF injections |  | 85-year-old, monthly anti-VEGF injections |  | One-time anti-VEGF injection: 75-year-old |
| --- | --- | --- | --- | --- | --- | --- | --- |
|  | optimistic assumptions | pessimistic assumptions | optimistic assumptions | pessimistic assumptions | optimistic assumptions | pessimistic assumptions | pessimistic assumptions |
| 1/10,000 | 0.003% | 0.027% | 0.006% | 0.054% | 0.013% | 0.109% | 0.009% |
| 1/1,000 | 0.032% | 0.274% | 0.063% | 0.544% | 0.127% | 1.09% | 0.091% |
| 1/100 | 0.319% | 2.74% | 0.631% | 5.44% | 1.27% | 10.9% | 0.906% |
| 1/10 | 3.19% | 27.4% | 6.31% | 54.4% | 12.6% | Likely harm | 9.06% |

For example, for a one-time procedure under the assumptions of the ‘pessimistic’ relevant COVID-19 attack rate of 43%, where full PPE is available, and the community prevalence of active COVID-19 is only one case per 10,000 population, then the procedure might be considered for a 75 year old if the likelihood of progression to severe visual impairment (or blindness) was as low as 0.001% or more (see Table 2, final column). For the same scenario, if PPE is not available and 10% of the population have active COVID-19 then the likelihood of progression to permanent severe visual impairment without treatment would need to be greater than 9.06% (see Table 3, final column).

Repeated treatments (eg monthly injections for six months) expose the patient and healthcare staff to a much higher chance of spreading the infection and results reflect that fact.

Results are rounded to 3 significant figures; Optimistic assumptions: 5% attack rate in healthcare setting, 10% of COVID-19 cases have protracted illness; Pessimistic assumptions: 43% attack rate in healthcare setting, 35% of COVID-19 cases have protracted illness; Monthly injections: anti-VEGF injection once per month for 6-month period.

Results are rounded to 3 significant figures; Optimistic assumptions: 5% attack rate in healthcare setting, 10% of COVID-19 cases have protracted illness; Pessimistic assumptions: 43% attack rate in healthcare setting, 35% of COVID-19 cases have protracted illness; Monthly injections: anti-VEGF injection once per month for 6-month period.

Table 4 displays the expected HALY benefit to the individual patient having treatment. It reflects the expected HALY loss from vision deterioration minus the expected impact on the patient from COVID-19, in the situation where the probability of progression to visual loss is 5%.

**Table 4:** Expected net HALY benefit to patient assuming a 5% probability of progression from moderate to severe vision loss

| Active community COVID-19 case prevalence | 65-year-old, monthly anti-VEGF injections (optimistic assumptions) | No PPE: 65-year-old, monthly anti-VEGF injections, (optimistic assumptions) | 85-year-old, monthly anti-VEGF injections (pessimistic assumptions) | No PPE: 85-year-old, monthly anti-VEGF injections, (pessimistic assumptions) |
| --- | --- | --- | --- | --- |
| 1/10,000 | 0.122 | 0.122 | 0.032 | 0.032 |
| 1/1,000 | 0.122 | 0.121 | 0.032 | 0.029 |
| 1/100 | 0.122 | 0.118 | 0.031 | 0.002 |
| 1/10 | 0.120 | 0.090 | 0.020 | -0.273 |

Optimistic assumptions: 5% attack rate in healthcare setting, 10% of COVID-19 cases have protracted illness; Pessimistic assumptions: 43% attack rate in healthcare setting, 35% of COVID-19 cases have protracted illness; Monthly injections: anti-VEGF injection once per month for 6-month period; HALY: health adjusted life-years; PPE: personal protective equipment

For example, where full PPE is not available, the attack rate is 43% (pessimistic assumptions), and the patient is aged 85 years needing monthly treatments, then results for this scenario indicate the expected HALY benefit (accounting for COVID-19 impact and vision loss avoided) is +0.032 at a COVID-19 prevalence of one per 10,000, but becomes an expected loss of -0.273 HALYs if one in 10 of the population are infected.

Applying the assumption that correct use of good quality PPE reduces the likelihood of transmission by 96%, we found HALY benefits in all scenarios where the probability of progression to severe visual impairment without treatment was at least 4.36% (Table 2).

## Discussion

Our results indicate that for the most plausible community prevalence rates of COVID-19 and with availability of appropriate PPE, then ophthalmic procedures should be considered more beneficial than the likely harm from COVID-19 in many cases provided there is a non-negligible probability of progression to severe visual impairment in the better seeing eye without treatment. The reason for this result is easily illustrated by considering that the HALYs lost when someone suffers severe visual impairment for 5 years are equivalent to nearly 400 moderate cases of infectious disease lasting 2 weeks each.^15^ The difference in disease burden expressed as annualised disability weight (DW) for a shift from moderate distance vision impairment to severe distance vision impairment is 0.153, whereas the DW for a ‘moderate infectious disease’ is 0.051, which only applies for the duration of illness.^15^ COVID-19 can be a severe and deadly disease for a minority of patients, nevertheless the burden of vision loss is significant and ongoing.

Notably, for all scenarios where COVID-19 prevalence is less than one per 100 population and proper PPE is available, a 1% likelihood of progression to severe visual impairment or blindness without treatment will tend to justify treatment. However, if there was a rapid escalation of COVID-19 cases, the ‘break even’ point at which HALYs lost rather than gained from providing care will change. We find that if one in 10 members of the population carries active virus, and PPE is in short supply, then the expected harms from COVID-19 outweigh the benefits of anti-VEGF treatments for older patients even when the chance of severe visual impairment without treatment is very high.

A number of other factors need to be considered beyond our simple model. Firstly, we encourage the development of more sophisticated and accurate epidemiological models of the likelihood of COVID-19 transmission in healthcare settings, given a range of contextual factors. We also encourage the development of estimates of the probability of severe visual impairment if eye conditions are untreated. This would allow models to output the expected HALYs gained and lost, rather than the probability of progression (requiring clinical judgement) at which the intervention ‘breaks even’ in terms of HALYs.

Secondly, given the high number of undetected COVID-19 cases in the community then ideally the true prevalence is established by random testing. In Austria, as at early April 2020, such studies revealed an active case prevalence of 0.33%.^25^ Early random community testing in New Zealand indicated zero community prevalence as at 20 April, 2020.^26^ However, an MIT modelling study concluded that the true number of cases in communities could be an order of magnitude higher than confirmed cases.^27^ This needs to be considered in any application of our approach.

Thirdly, our evaluation considers individual cases and assumes that the healthcare team is different each time for repeated procedures. This means that cumulative risk to individual health care workers who may perform many procedures, is not evaluated. The risk to individuals who are likely to be repeatedly exposed ought to be considered when weighing benefits and harms. That said, even without effective PPE, and with a COVID-19 population prevalence of one per 10,000, the expected HALY loss to health care workers due to COVID-19 infection appears to be approximately -0.00002 per person per procedure. The reason is that for the vast majority of people COVID-19 is a mild infectious illness for a week or two.^28^ However, consideration might be given to a policy of halting procedures on the basis of health care team safety, irrespective of risk of blindness, where community prevalence rates are very high and PPE is not available. For example, at 10% COVID-19 prevalence, without available PPE, the net probable impact on healthcare workers is -0.022 HALYs per staff member per procedure. This may be considered unacceptable, irrespective of benefit to patients.

Finally, local COVID-19 mitigation strategies should be considered. In contexts where mitigation of disease impact or suppression of the number of cases is the goal, then the arguments above will apply. However, if the aim is elimination of COVID-19 then extra consideration might be given to avoiding all personal contact, including ophthalmologic procedures. However, if patients recover at home in quarantine, and it is possible to keep health care teams in small ‘bubbles’, then the risks might be managed.

A further consideration when weighing up whether to proceed with interventions or not is future demand. For example, without continuing elective anti-VEGF injections where feasible, there is a risk of post-pandemic surge in demand. Health authorities need to plan ahead in order to avoid a mismanaged flow, and minimize staff burnout.^29^ We are mindful that the reasoning described in this paper will apply to a number of other procedures where untreated conditions are likely to result in a high burden of disease. The benefits and harms of performing procedures for these conditions should also be quantified.

The risk of COVID-19 spread can be further mitigated by appropriate health facility processes, including staff rotation, physical distancing, handwashing and disinfection, alert signs, no touch payments and many other measures.^3^ A study of 30 confirmed and suspected COVID-19 patients undergoing caesarean section reported no infection of healthcare workers.^30^ This suggests that transmission can be avoided under optimal conditions. Effective PPE makes a dramatic difference to the calculus. Furthermore, patients and staff could be tested for COVID-19 and could remain in self-quarantine following contact.

During this unique period of a global pandemic, the equation for continuing to provide care becomes more complicated. For ophthalmologists, there is a mindset change from patient-centered (preventing vision loss) to a community-centered (preventing spread of infectious disease and death) perspective. The personal health risks that the ophthalmology team and our patients take must be carefully considered. The average age for nAMD patients increases their risk for severe illness and death given age is one of the most important risk factors.^31^ Likewise, ophthalmologists are currently overrepresented in physician deaths attributed to COVID-19 (4% of all physicians) possibly reflecting their close contact with patients. Dr. Li Wenliang, the ophthalmologist and whistleblower who informed the world of COVID-19, died of the disease.^32^ Our calculations show that with appropriate PPE (sufficient to reduce transmission probability by 96%), the transmission risks are almost zero, even with moderately high COVID-19 prevalence, however the limiting factor will be the availability of this equipment.

In locations where appropriate PPE is not available then decisions might be guided by Table 3, which shows that there is net benefit in proceeding with treatments for younger patients with even low probability of eye disease progression, whereas older patients would need to be somewhat likely (75 years) or very likely (85 years) to progress for the benefits of treatment to outweigh the harms of COVID-19. This is particularly so when the attack rate tends towards our pessimistic assumption of 43% and/or many patients suffer lingering symptoms.

## Limitations

This analysis is only a proof-of-concept using a simple coarse-grained model. We have not accounted for a number of variables including the possible spread of any COVID-19 acquired during procedures to others beyond the patient and healthcare team. For repeat procedures across a six-month pandemic period we have assumed a constant community prevalence of COVID-19, however this may peak and decline. More sophisticated epidemiological models should be developed to account for these variables. Also, our model applies the DW for progression from ‘moderate visual impairment’ to ‘severe visual impairment/blindness’ as an immediate transition, which does not account for slow deterioration and will likely overestimate HALY losses. We have also assumed just three healthcare staff exposed to each other and each patient, however, real clinical interactions may involve more close contacts than this. We have assumed that high-quality PPE reduces the chance of infection by 96% (ie one in twenty-five close contacts using PPE are exposed to the risk of infection). Although, this could still be conservative, as we note above, some reports suggest very low or no COVID-19 transmission in surgical settings (zero infections during 30 caesarean deliveries in one study). Our analysis also does not consider whether health resources might better be spent treating COVID-19 patients. Our analysis was performed in the context of New Zealand’s population demographic, including age at treatment and health adjusted life-expectancy and findings may not generalise to other settings. Finally, the COVID-19 pandemic is evolving and so the parameters used in the model are dynamic and subject to change over time as more information and further research becomes available.

## Conclusion

Recent clinical guidelines emphasise the need for clinicians to consider the benefits and harms of performing clinical procedures during the COVID-19 pandemic. We estimated the expected HALYs lost due to COVID-19 transmission in the clinical setting, and the expected HALY gains from preventing progression of better seeing eye disease from moderate impairment to severe impairment.

Our simple calculations suggest that with appropriate PPE, the threshold for carrying out interventions is likely to be low, with a 1% chance of severe visual impairment or blindness without treatment justifying interventions in most scenarios we analysed, except where there is very high population prevalence of COVID-19 disease or patients are older.

## Data Availability

Mathematical modelling described in methods section.

## REFERENCES

1. Fedarazione Nazionale degli Ordini del Medici Chirughi e degli Odontoiatri. Elenco dei Medici caduti nel corso dell ‘epidemia di Covid-19. Rome: Federazione Nazionale degli Ordini dei Medici Chirurghi e degli Odontoiatri; 2020. Accessed April 2020. Available from: https://portale.fnomceo.it/elenco-dei-medici-caduti-nel-corso-dellepidemia-di-covid-19/.

2. Breazzano MP, Shen J, Abdelhakim AH, Glass LRD, Horowitz JD, Xie SX, et al. Resident physician exposure to novel coronavirus (2019-nCoV, SARS-CoV-2) within New York City during exponential phase of COVID-19 pandemic: Report of the New York City Residency Program Directors COVID-19 Research Group. *MedRxiv*. 2020. doi: https://doi.org/10.1101/2020.04.23.20074310.

3. Gharebaghi R, Desuatels J, Moshirfar M, Parvizi M, Daryabari S, Heidary F. COVID-19: Preliminary Clinical Guidelines for Ophthalmology Practices. Med Hypothesis Discov Innov Ophthalmol. 2020;9:149–58. doi: 10.1136/bmjophth-2020-000487.

4. The Lancet. COVID-19: protecting health-care workers. Lancet. 2020;395:922. doi: 10.1016/S0140-6736(20)30644-9.

5. Lau PE, Jenkins KS, Layton CJ. Current Evidence for the Prevention of Endophthalmitis in Anti-VEGF Intravitreal Injections. J Ophthalmol. 2018;2018:8567912. doi: 10.1155/2018/8567912.

6. Parikh R, Ross RS, Sangaralingham LR, Adelman RA, Shah ND, Barkmeier AJ. Trends of Anti-Vascular Endothelial Growth Factor Use in Ophthalmology Among Privately Insured and Medicare Advantage Patients. Ophthalmol. 2016;124:352–8. doi: 10.1016/j.ophtha.2016.10.036.

7. Ernst & Young. Age-related Macular Degeneration Model of care assessment and recommendations. Ernst & Young; 2017.

8. Royal Australian and New Zealand College of Ophthalmologists (RANZCO). RANZCO New Zealand Branch Triage Guidelines - COVID-19 Level 4. Sydney: Royal Australian and New Zealand College of Ophthalmologists; 2020. Accessed April 2020. Available from: https://ranzco.edu/wp-content/uploads/2020/04/RANZCO-triage-NZ-modification-level-4.pdf.

9. Seddon J, Khurana R. Coronavirus and Your Macular Degeneration Care. Northhampton: American Macular Degeneration Foundation; 2020. Accessed April 2020. Available from: https://www.macular.org/2020/03/19/coronavirus-and-your-macular-degeneration-care.

10. Ozturk M, Harris ML, Nguyen V, Barthelmes D, Gillies MC, Mehta H. Real-world visual outcomes in patients with neovascular age-related macular degeneration receiving aflibercept at fixed intervals as per UK licence. Clin Exp Ophthalmol. 2018;46:407–11. doi: 10.1111/ceo.13085.

11. Gale RP, Mahmood S, Devonport H, Patel PJ, Ross AH, Walters G, et al. Action on neovascular age-related macular degeneration (nAMD): recommendations for management and service provision in the UK hospital eye service. Eye (Lond). 2019;33:1–21. doi: 10.1038/s41433-018-0300-3.

12. Nguyen V, Vaze A, Fraser-Bell S, Arnold J, Essex RW, Barthelmes D, et al. Outcomes of Suspending VEGF Inhibitors for Neovascular Age-Related Macular Degeneration When Lesions Have Been Inactive for 3 Months. Ophthalmol Retina. 2019;3:623–8. doi: 10.1016/j.oret.2019.05.013.

13. Buckle M, Lee A, Mohamed Q, Fletcher E, Sallam A, Healy R, et al. Prevalence and incidence of blindness and other degrees of sight impairment in patients treated for neovascular age-related macular degeneration in a well-defined region of the United Kingdom. Eye (Lond). 2015;29:403–8. doi: 10.1038/eye.2014.296.

14. Thomas DS, Warwick A, Olvera-Barrios A, Egan C, Schwartz R, Patra S, et al. Estimating excess visual loss in people with neovascular age-related macular degeneration during the COVID-19 pandemic. *MedRxiv*. 2020. doi: https://doi.org/10.1101/2020.06.02.20120642.

15. Salomon JA, Haagsma JA, Davis A, de Noordhout CM, Polinder S, Havelaar AH, et al. Disability weights for the Global Burden of Disease 2013 study. Lancet Glob Health. 2015;3:e712-23. doi: 10.1016/s2214-109x(15)00069-8.

16. Greenhalgh T, Knight M, A’Court C, Buxton M, Husain L. Management of post-acute covid-19 in primary care. BMJ. 2020;370:m3026. doi: 10.1136/bmj.m3026.

17. Tenforde WM, Kim SS, Lindsell CJ, Rose EB, Shapiro NI, Files DC, et al. Symptom Duration and Risk Factors for Delayed Return to Usual Health Among Outpatients with COVID-19 in a Multistate Health Care Systems Network — United States, March–June 2020. MMWR Morb Mortal Wkly Rep. 2020;69:993–8. doi: http://dx.doi.org/10.15585/mmwr.mm6930e1.

18. WHO-China Joint Mission. Report of the WHO-China Joint Mission on Coronavirus Disease 2019 (COVID-19). World Health Organization; 2020.

19. Park S, Kim Y, Yi S, Lee S, Na B, Kim C. Coronavirus disease outbreak in call center, South Korea. Emerg Infect Dis. 2020;26. doi: 10.3201/eid2608.

20. Immigration New Zealand. Building and keeping a health workforce. Wellington: Immigration New Zealand; 2015. Accessed April 2020. Available from: https://www.immigration.govt.nz/about-us/media-centre/newsletters/settlement-actionz/actionz3/building-and-keeping-a-health-workforce.

21. Kvizhinadze G, Wilson N, Nair N, McLeod M, Blakley T. How much can society spend on life-saving interventions at different ages while remaining cost effective? Estimates using New Zealand health system costs, morbidity, and mortality data. Population Health Metrics. 2015;13. doi: 10.1186/s12963-015-0052-2.

22. Chu DK, Akl EA, Duda S, Solo K, Yaacoub S, Schiinemann HJ. Physical distancing, face masks, and eye protection to prevent person-to-person transmission of SARS-CoV-2 and COVID-19: a systematic review and meta-analysis. Lancet. 2020. doi: https://doi.org/10.1016/S0140-6736(20)31142-9.

23. Verity R, Okell L, Doriagatti I, Winskill P, Whittaker C, Imai N. Estimates of the severity of COVID-19 disease. *MedRxiv*. 2020. doi: https://doi.org/10.1101/2020.03.09.20033357.

24. Ferguson N, Laydon D, Nedjati-Gilani G, Imai N, Ainslie K, Baguelin M, et al. Impact assessment of non-pharmaceutical interventions (NPIs) to reduce COVID-19 mortality and healthcare demand. London: Imperial College; 2020. Report No.: 9. doi: https://doi.org/10.25561/77482.

25. Ogris G, Hofinger C. COVID-19 Prevalence: Media information, April 10. Sora Institut; 2020.

26. New Zealand Government. Transcript of COVID-19 media conference - 20 April. Wellington: New Zealand Government; 2020. Accessed April 2020. Available from: https://covid19.govt.nz/latest-updates/covid-19-media-conference-20-april/transcript-of-covid-19-media-conference-20-april/.

27. Rahmandad H, Lim TY, Sterman J. Estimating COVID-19 Under-Reporting Across 86 Nations: Implications for Projections and Control. *SSRN*. 2020. doi: https://ssrn.com/abstract=3635047.

28. Sharma R, Agarwal M, Gupta M, Somendra S, Saxena SK. Clinical Characteristics and Differential Clinical Diagnosis of Novel Coronavirus Disease 2019 (COVID-19). Coronavirus Disease 2019. 2020:55-70. doi: 10.1007/978-981-15-4814-7_6.

29. Zhang H, Bo L, Lin Y, Li F, Sun S. Response of Chinese Anesthesiologists to the COVID-19 Outbreak. Anesthesiology. 2020. doi: 10.1097/ALN.0000000000003300.

30. Yue L, Han L, Li Q, Zhong M, Wang J, Wan Z, et al. Anaesthesia and infection control in cesarean section of pregnant women with coronavirus disease 2019 (COVID-19). *MedRxiv*. 2020. doi: https://doi.org/10.1101/2020.03.23.20040394.

31. Jordan R, Adab P, Cheng K. Covid-19: risk factors for severe disease and death. BMJ. 2020;368. doi: 10.1136/bmj.m1198.

32. Ing E, Xu A, Salimi A, Tourun N. Physician Deaths from Corona Virus Disease (COVID-19). *MedRxiv*. 2020. doi: https://doi.org/10.1101/2020.04.05.20054494.

